# Combination of Spironolactone and Sitagliptin Improves Clinical Outcomes of Outpatients with COVID-19: A Prospective Cohort Study

**DOI:** 10.1101/2022.01.21.22269322

**Authors:** Mohammad Ali Davarpanah, Reuben Adatorwovor, Yasaman Mansoori, Fatemeh Sadat Rajaie Ramsheh, Amir Parsa, Mehdi Hajiani, Hossein Faramarzi, Ramakanth Kavuluru, Kamyar Asadipooya

## Abstract

**Rationale:** Coronavirus disease 2019 (COVID-19) leads to hospitalization and death, especially in elderly and those with comorbidities. There are evidences showing that sitagliptin and spironolactone can potentially improve the clinical outcomes of COVID-19 cases.

**Objective:** In this observational study on acutely symptomatic outpatient COVID-19 cases, we investigated the effects of spironolactone and sitagliptin on the outcomes of the disease.

**Methods:** This prospective cohort study was conducted at Shiraz University of Medical Sciences Clinics during the fifth wave of the COVID-19 pandemic between July 2021 and September 2021. We followed mild to moderate symptomatic COVID-19 patients, who were treated with either combination (spironolactone 100 mg daily and sitagliptin 100 mg daily) or standard (steroid, antiviral and/or supportive care) therapy up to 30 days. Our primary outcome was hospitalization rate. The secondary outcomes included ER visit, duration of disease, and complications, such as hypoglycemia, low blood pressure or altered mental status.

**Results:** Of the 206 patients referred to clinics, 103 received standard therapy and 103 treated with combination therapy. There were no significant differences in baseline characteristics, except for slightly higher clinical score in control group (6.92 ± 4.01 control, 4.87 ± 2.92 combination; *P <0.0001*). Treatment with combination therapy was associated with lower admission rate (5.8% combination, 22.3% control; *P = 0.0011*), ER visits (7.8% combination, 23.3% control; *P = 0.0021*) and average duration of symptoms (6.67 ± 2.30 days combination, 18.71 ± 6.49 days control; *P =<0.0001*).

**Conclusion:** In this prospective cohort study of acutely ill outpatients with COVID-19, the combination of sitagliptin and spironolactone reduced duration of COVID infection and hospital visits better than standard therapeutic approaches. The effects of combination of sitagliptin and spironolactone in COVID-19 patients should be further verified in a double blind, randomized, placebo-controlled trial.

**Iranian Registry of Clinical Trials:** IRCT registration number: IRCT20201003048904N2, Registration date: December 10, 2020.

## Introduction

More than five million people have died from coronavirus disease 2019 (COVID-19) since the pandemic has started (1). Patients with comorbidities, especially men and elderly, are at higher risk for death or hospitalization (2). Although being vaccinated was associated with reduced hospitalization and mortality (3), there are concerns regarding vaccine efficacy against new variants of coronavirus (4). Therefore, the role of an effective and safe treatment would be crucial.

There are substantial variations between different guidelines, possibly due to lack of enough evidence or limited efficacy of current treatments (5). Monoclonal antibodies, especially the combination form and early treatment, improved the outcomes by reducing duration of illness, hospitalization and death (6,7). However, severe acute respiratory syndrome coronavirus 2 (SARS-CoV-2) has rapidly evolved with the mutations of the spike protein and generation of new variants (8,9), which ended up reducing the beneficial effects of vaccine and monoclonal antibodies (10,11). In addition, some of the therapeutic options, such as monoclonal antibodies and convalescent plasma, are not broadly available and generally require a hospital visit. Hence, blocking virus entry into the cells through its receptor would be a potential alternative to reduce viral replication and severity of the disease.

SARS-CoV-2 enters into the host cell mainly through interacting with the transmembrane protein angiotensin-converting enzyme 2 (ACE2) (12) and, probably, dipeptidyl peptidase 4 (DPP4) (13,14). SARS-CoV-2 has even higher affinity to human ACE2 compared to SARS-CoV (15), which makes it dependent on ACE2 as a major receptor for cell entry (16). Nevertheless, during the acute infection, the enormous virus replication may overwhelm the ACE2 receptors and undertake DPP4 receptors for cellular entry. As a result, blocking virus entry during the acute illness by reducing the interaction of SARS-CoV-2 spike glycoprotein with ACE2 and DPP4 receptors simultaneously can result in better clinical outcomes.

The transmembrane protease serine protease-2 (TMPRSS-2) and ADAM metallopeptidase domain 17 (ADAM17) participate in cleaving ACE2 and priming the S glycoprotein, which potentiate the virus endocytosis. ACE2 has two forms: soluble and membrane bound (16-18). The binding of SARS-CoV-2 to soluble ACE2 increases weight and diameter of viral particles, leading to a higher chance of virus attachment to the cell membrane and engulfment (19). Spironolactone, as a mineralocorticoid receptor blocker with anti-androgenic effects, can reduce TMPRSS-2 expression (20,21), inhibit ADAM17 (22), decrease soluble ACE2 plasma level (23), but increases ACE2 expression on cell membrane (24). Moreover, spironolactone reduces inflammation with the advantageous effects on oxidative injury and hypercoagulability (21). Consequently, spironolactone can potentially reduce viral entry and improve clinical conditions of acutely ill COVID-19 cases.

DPP4 inhibitors are associated with lower mortality and intubation risk in COVID-19 patients with diabetes (25-27). Moreover, they could improve clinical outcomes and inflammatory markers (28). This could be potentially due to their immunomodulatory roles (29) and blocking the possible interaction between SARS-CoV-2 spike protein and DPP4 receptors (13,30).

In this study, we propose and assess the hypothesis that spironolactone and sitagliptin (a DPP4 inhibitor), by affecting inflammation and coronavirus cell entry through ACE2 and DPP4, could potentially reduce hospitalization of outpatients with COVID-19, without causing serious adverse events.

## Methods

### Study design and population

A prospective cohort study was conducted at Shiraz University of Medical Sciences (SUMS) clinics during the fifth wave of the COVID-19 pandemic in Iran, when the delta and subsequent variants of SARS-CoV-2 were dominant (31). The date range for participant recruitment and follow-up was between August 2, 2021 and October 22, 2021. The attending physicians provided the care in the institutionalized clinics (Motahhari Clinic, Shiraz University of Medical Sciences). We enrolled 216 patients, but 10 patients were lost at follow-up (**Figure 1**). Thus, we included 206 confirmed or probable adult outpatient cases of SARS-CoV-2 infection according to WHO and CDC definitions (32). The eligible patients were at least 18 years of age and had laboratory confirmed SARS-CoV-2 infection (nasal/throat swabs positive for SARS-CoV-2 by RT-PCR) or positive history of exposure to COVID-19 patients besides typical clinical manifestations (33). Exclusion criteria were ongoing mineralocorticoid receptor antagonists (including spironolactone) or DPP4 inhibitors (including sitagliptin) treatment. There were two active clinics (clinics A and B). The outside physicians, who are not aware of the study, were referring the patients to each clinic. The attending physicians enrolled all referred patients into the study if they were eligible, not excluded, or did not refuse taking medications (the spironolactone and sitagliptin intervention). The investigators had no influence on selection of the patients. Patients were recruited into two cohort groups (103 patients either in cohort group A or B after excluding the patients who lost follow up). Patients in cohort group A received the standard therapy for COVID-19, including supportive care (oral hydration, antipyretics, etc.), antivirals (remdesivir or favipiravir), or steroids based on the severity of their symptoms. The steroid was given during the second week visit or after with worsening of symptoms or oxygen saturation less than 90% despite supportive care according to the national guideline, specifically, dexamethasone 8 mg daily for 3 days, then the average dose of prednisolone 25 mg daily for 4 days (starting with 50 mg daily, then tapered and discontinued in 4 days). Patients in cohort group B received spironolactone 100 mg daily and sitagliptin 100 mg daily with or without antiviral therapy. We prescribed a 10-day regimen of combination and told the patients to continue the medications until recovery or call the clinic for further refills. We used combination therapy as an add-on to the antiviral (favipiravir) for 22 patients at the beginning of study according to the national COVID-19 committee recommendations (34). We did not use any steroids in the combination group.

**Figure 1.**
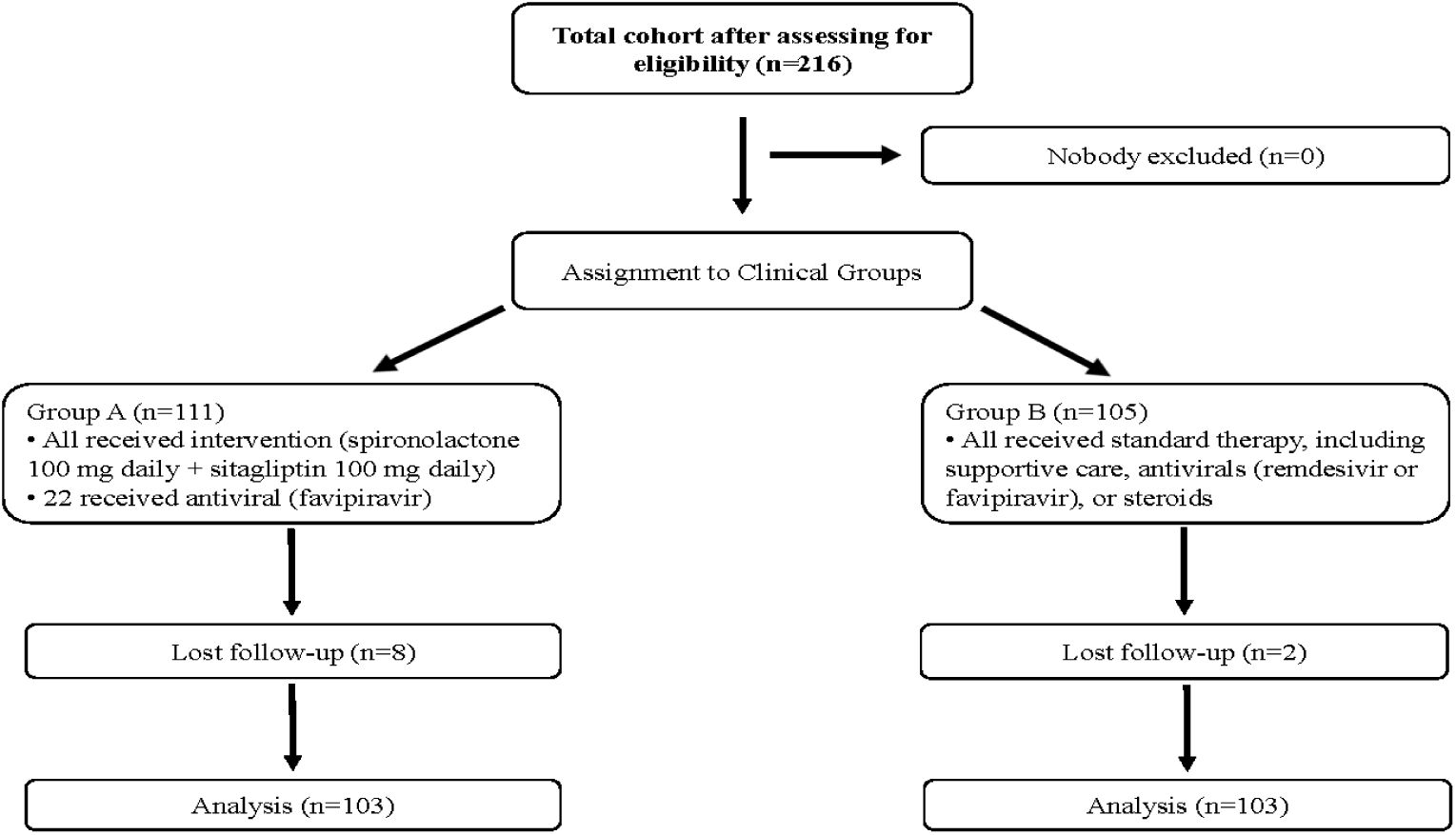
Flow chart of the cohort study design

We calculated the clinical severity score (**Table 1**) determined by sex, age, clinical conditions, functional performance, and oxygen requirement using a previously validated scoring tool for outcomes prediction in acutely ill adult patients with COVID-19. The scale ranged from 0 to 29, with higher scores reflecting worse clinical conditions and scores greater than four showing high sensitivity for recognizing patients with high risk for adverse events (**Table 1**) (35).

**Table 1.** OPD clinical severity score determines the severity of the disease based on clinical conditions, functional performance and oxygen requirement. The raising scores means worse clinical status (35).

| *Variable* | *Range* |
| --- | --- |
| *Respiratory rate per minute* | 0-3 |
| *Oxygen saturation %* | 0-3 |
| *Heart rate per minute* | 0-3 |
| *Systolic blood pressure mmHg* | 0-3 |
| *Temperature* | 0-3 |
| *Alertness* | 0 and 3 |
| *Inspired oxygen* | 0 and 2 |
| *Sex* | 0 and 1 |
| *Age (years)* | 0-4 |
| *Functional performance (activity)* | 0-4 |
| *Total score* | 29 |

| **Variable** | **Range** | **Score** |
| --- | --- | --- |
| Sex | Female | 0 |
|  | Male | 1 |
| Respiratory rate per minute | 12-20 | 0 |
|  | 9-11 | 1 |
|  | 21-24 | 2 |
|  | <9 or >24 | 3 |
| Oxygen saturation (%) | >95 | 0 |
|  | 94-95 | 1 |
|  | 92-93 | 2 |
|  | <92 | 3 |
| Heart rate per minute | 51-90 | 0 |
|  | 41-50 or 91-110 | 1 |
|  | 111-130 | 2 |
|  | <41 or >130 | 3 |
| Systolic blood pressure mmHg | 111-219 | 0 |
|  | 101-110 | 1 |
|  | 91-100 | 2 |
|  | <91 or >219 | 3 |
| Temperature °C | 36.1-38.0 | 0 |
|  | 35.1-36.0 or 38.1-39.0 | 1 |
|  | >39.0 | 2 |
|  | <35.1 | 3 |
| Alertness | Alert | 0 |
|  | Confused or disoriented | 3 |
| Oxygen requirement | No nasal oxygen | 0 |
|  | Nasal oxygen | 2 |
| Age per years | 18-49 | 0 |
|  | 50-65 | 2 |
|  | 66-80 | 3 |
|  | >80 | 4 |
| Functional performance | Normal activity, no restriction | 0 |
|  | Mild to moderate limitation | 1 |
|  | Limited activity and self-care | 2 |
|  | Limited to self-care | 3 |
|  | Bed ridden | 4 |
| Total score |  | 29 |

Two follow-ups were conducted one week and one month after the intervention in person or by phone calls. We asked the patients about the need for hospitalization and ER visits. We also recorded duration of disease, adherence to the medication, medication intolerance, and possible complications, such as hypotension, hypoglycemia or drug reaction.

Recovery of COVID-19 was defined as a resolution of fever without using fever-reducing agents and improvement of other symptoms. We compared the clinical outcomes, including hospitalization, emergency room (ER) visits, duration of treatment and complications between the groups.

### Clinical data

The research physicians collected and recorded data, including the baseline characteristics, medical history (medications, comorbidities), clinical conditions, hospitalization rate, ER visits, and complications. The comorbidities include diabetes (on medication or hemoglobin A1C > 6.5), hypertension (on medication or blood pressure > 140/90), cardiovascular disease, renal disease, liver disease, lung disease, neurologic disorder, malignancy, hematologic disorders, immune deficiency (transplant etc.), rheumatologic disorder, and hypothyroidism.

The primary endpoint was the rate of hospitalization between two groups. Secondary endpoints included emergency room (ER) visits, duration of treatment, and intervention related complications.

The research physicians had routinely provided a verbal informed consent at the time of data collection. They had explained the purpose of study, benefits and risks of medications and the process of data collection and analysis. The ethics committee of Shiraz University of Medical Sciences (IR.SUMS.MED.REC.1399.550) approved the study. We designed the study according to the declaration of Helsinki and Iranian national guidelines for ethics in research. The research physicians de-identified the patients’ information after collection and prepared a code for re-identification. The University of Kentucky received the de-identified data for statistical analysis. The Shiraz University of Medical Sciences sponsored the study. The Institutional Review Boards of the University of Kentucky, and Shiraz University of Medical Sciences approved the study. The current trial is registered as an addendum to the previous trial, which was conducted on the admitted patients (36). The authors confirm that all ongoing and related trials for these drugs are registered.

## Statistical Analysis

Descriptive summaries for the data from the two cohort groups were reported in terms of means and standard deviations, and counts and their corresponding proportions. Specifically, we compared the baseline characteristics across the cohort groups using t- and Mantel-Haenzel chi-square tests for continuous and categorical variables respectively. The main outcome of this analysis is COVID-19 hospitalization and we tested hospitalization rate among the cohort groups using the Kaplan-Meier curve and log-rank test.

Additionally, we compare the odds of hospitalization using univariate logistic regression model for each covariate. Multiple logistic regression model was fitted to dichotomized hospitalization outcome while adjusting for all covariates in a multivariate setting. Such covariates include demographic variables such as age, sex, and clinical variables including duration of disease (COVID-19), ER visit, baseline clinical score, prior COVID-19 infection, vaccination status, and comorbidities. We defined comorbidities to be patients who had any of the following: diabetes, hypertension, heart disease, kidney disease, lung disease, liver disease, hypothyroidism, cancer, and hematologic disease. We presented the odds ratio, its 95% confidence interval with its associated p-value for each of the covariates in the model. The odds ratio compares the risk of hospitalization among the two cohort groups. A positive coefficient lower than 1 indicates lower risk while a positive coefficient larger than 1 indicates a higher risk associated with the covariate in comparing hospitalization rate in the cohort groups.

All models were fitted using SAS logistics procedures and all analyses were performed using SAS Version 9.4 (TS1M1 SAS Institute Inc., Cary, NC) and R statistical Software. We used the standard 5% significance level for testing our entire hypothesis with the rejection of the null hypothesis for p-values less than 0.05.

## Results

### Patient Characteristics

We assigned 216 eligible patients at the beginning, but 10 patients were lost during follow-up. A total of 103 patients were treated with combination therapy (spironolactone 100 mg daily and sitagliptin 100 mg daily), while 103 patients received the standard of care. Almost half of them had +PCR results (57.3% standard, 49.5% combination; *P = 0*.*2650*), and the rest had history of exposure to COVID-19 with typical clinical manifestations. Baseline demographic characteristics of the two groups are shown in **Table 2**. The mean age was almost similar (41.50 ± 12.19 control, 42.45 ± 13.11 combination, *P = 0*.*5941*) and there were no significant differences in male sex (41.7% control, 35.0% combination, *P = 0*.*3170*). Both cohort groups did not have major differences concerning other demographic or clinical characteristics, except for clinical score was higher in control group (6.92 ± 4.01 control, 4.87 ± 2.92 combination; *P <0*.*0001*). In addition, there were no differences in comorbidities and medications, except combination group did not receive steroid (**Table 2**).

**Table 2.**
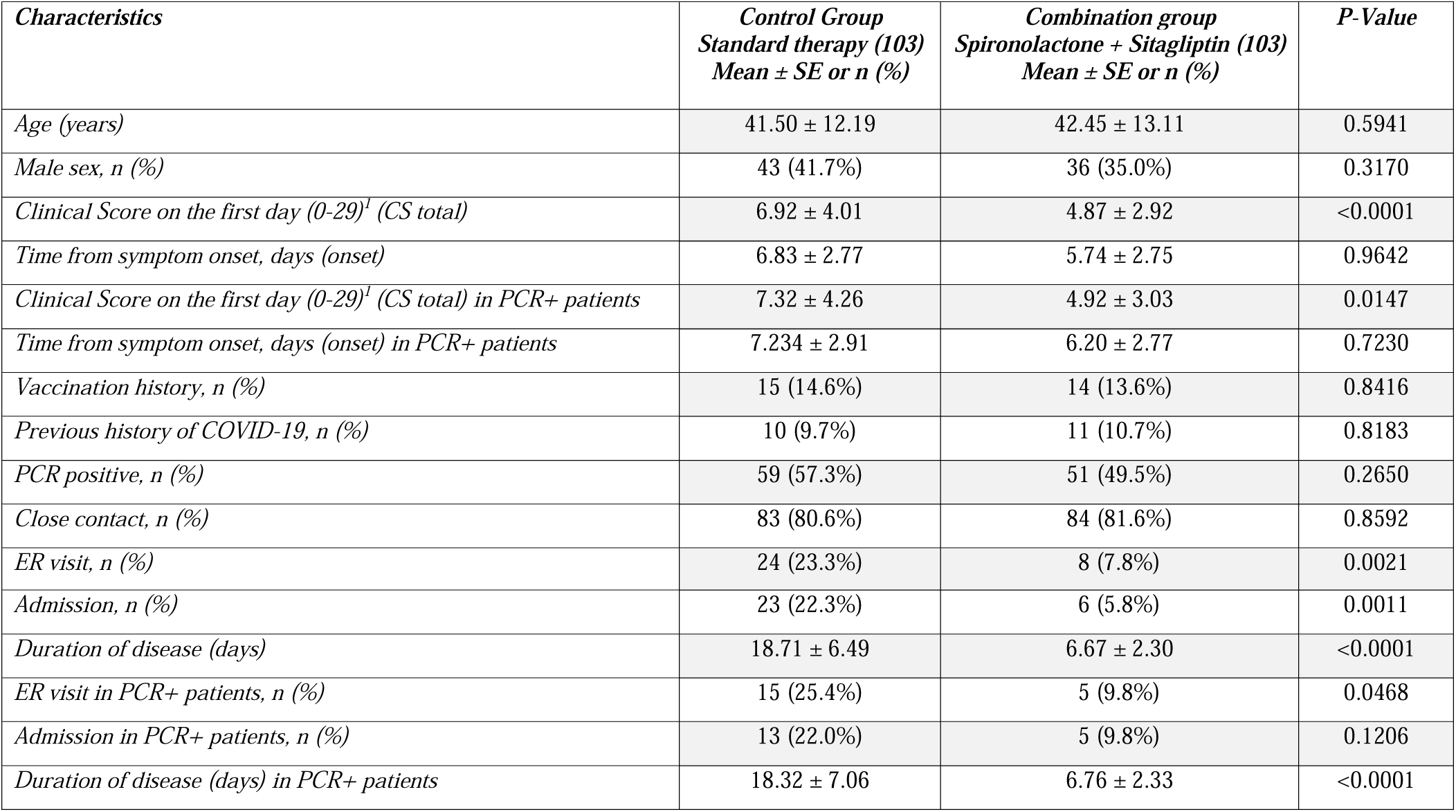
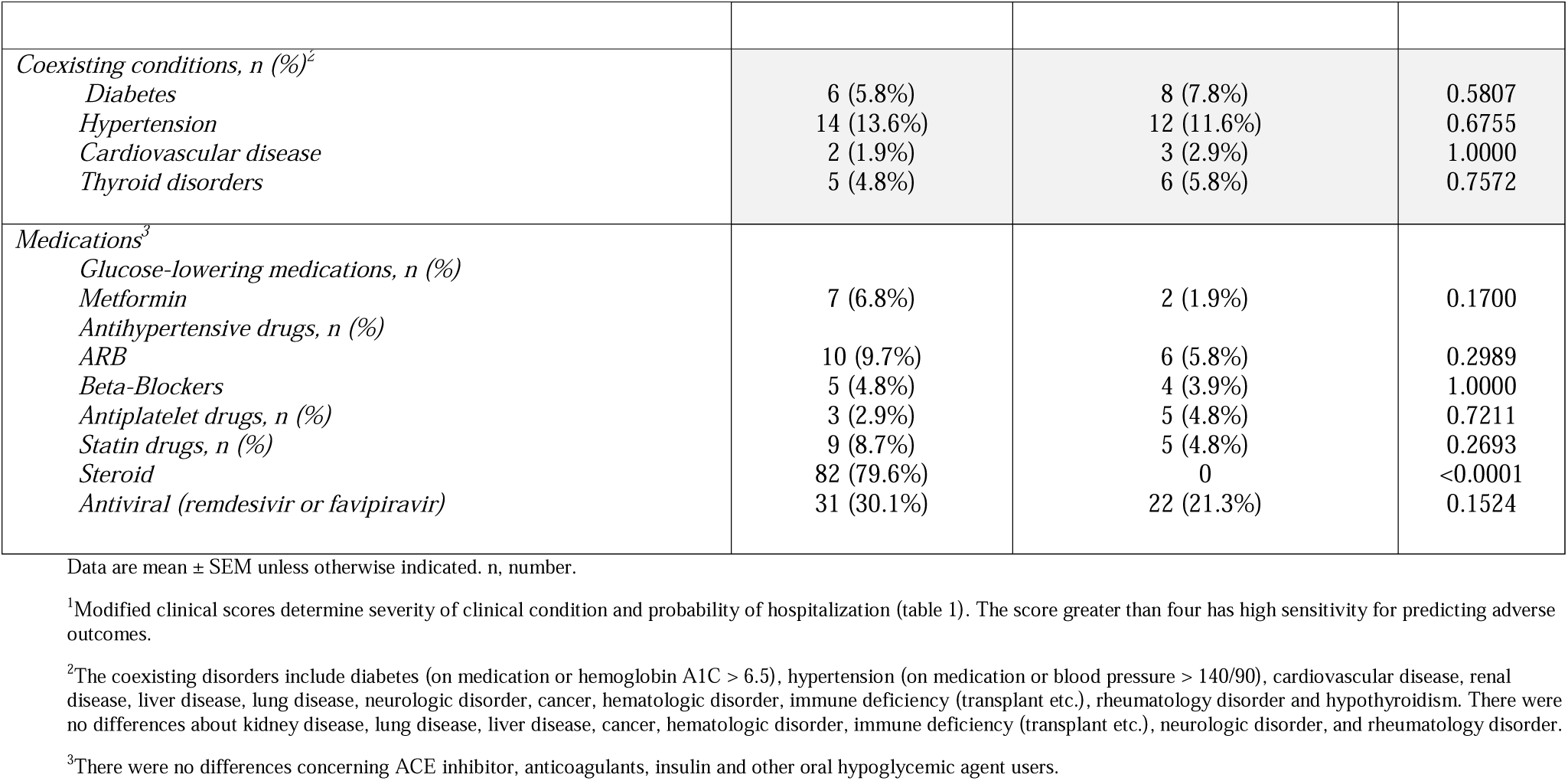
The baseline characteristics and outcomes of the study population

| *Characteristics* | *Control Group*<br>*Standard therapy (103)*<br>*Mean ± SE or n (%)* | *Combination group*<br>*Spironolactone + Sitagliptin (103)*<br>*Mean ± SE or n (%)* | *P-Value* |
| --- | --- | --- | --- |
| *Age (years)* | 41.50 ± 12.19 | 42.45 ± 13.11 | 0.5941 |
| *Male sex, n (%)* | 43 (41.7%) | 36 (35.0%) | 0.3170 |
| *Clinical Score on the first day (0-29)<sup>1</sup> (CS total)* | 6.92 ± 4.01 | 4.87 ± 2.92 | <0.0001 |
| *Time from symptom onset, days (onset)* | 6.83 ± 2.77 | 5.74 ± 2.75 | 0.9642 |
| *Clinical Score on the first day (0-29)<sup>1</sup> (CS total) in PCR+ patients* | 7.32 ± 4.26 | 4.92 ± 3.03 | 0.0147 |
| *Time from symptom onset, days (onset) in PCR+ patients* | 7.234 ± 2.91 | 6.20 ± 2.77 | 0.7230 |
| *Vaccination history, n (%)* | 15 (14.6%) | 14 (13.6%) | 0.8416 |
| *Previous history of COVID-19, n (%)* | 10 (9.7%) | 11 (10.7%) | 0.8183 |
| *PCR positive, n (%)* | 59 (57.3%) | 51 (49.5%) | 0.2650 |
| *Close contact, n (%)* | 83 (80.6%) | 84 (81.6%) | 0.8592 |
| *ER visit, n (%)* | 24 (23.3%) | 8 (7.8%) | 0.0021 |
| *Admission, n (%)* | 23 (22.3%) | 6 (5.8%) | 0.0011 |
| *Duration of disease (days)* | 18.71 ± 6.49 | 6.67 ± 2.30 | <0.0001 |
| *ER visit in PCR+ patients, n (%)* | 15 (25.4%) | 5 (9.8%) | 0.0468 |
| *Admission in PCR+ patients, n (%)* | 13 (22.0%) | 5 (9.8%) | 0.1206 |
| *Duration of disease (days) in PCR+ patients* | 18.32 ± 7.06 | 6.76 ± 2.33 | <0.0001 |
| *Coexisting conditions, n (%)*<sup>2</sup> |  |  |  |
| *Diabetes* | 6 (5.8%) | 8 (7.8%) | 0.5807 |
| *Hypertension* | 14 (13.6%) | 12 (11.6%) | 0.6755 |
| *Cardiovascular disease* | 2 (1.9%) | 3 (2.9%) | 1.0000 |
| *Thyroid disorders* | 5 (4.8%) | 6 (5.8%) | 0.7572 |
| *Medications*<sup>3</sup> |  |  |  |
| *Glucose-lowering medications, n (%)* |  |  |  |
| *Metformin* | 7 (6.8%) | 2 (1.9%) | 0.1700 |
| *Antihypertensive drugs, n (%)* |  |  |  |
| *ARB* | 10 (9.7%) | 6 (5.8%) | 0.2989 |
| *Beta-Blockers* | 5 (4.8%) | 4 (3.9%) | 1.0000 |
| *Antiplatelet drugs, n (%)* | 3 (2.9%) | 5 (4.8%) | 0.7211 |
| *Statin drugs, n (%)* | 9 (8.7%) | 5 (4.8%) | 0.2693 |
| *Steroid* | 82 (79.6%) | 0 | <0.0001 |
| *Antiviral (remdesivir or favipiravir)* | 31 (30.1%) | 22 (21.3%) | 0.1524 |

Data are mean ± SEM unless otherwise indicated. n, number.
<sup>1</sup>Modified clinical scores determine severity of clinical condition and probability of hospitalization (table 1). The score greater than four has high sensitivity for predicting adverse outcomes.
<sup>2</sup>The coexisting disorders include diabetes (on medication or hemoglobin A1C > 6.5), hypertension (on medication or blood pressure > 140/90), cardiovascular disease, renal disease, liver disease, lung disease, neurologic disorder, cancer, hematologic disorder, immune deficiency (transplant etc.), rheumatology disorder and hypothyroidism. There were no differences about kidney disease, lung disease, liver disease, cancer, hematologic disorder, immune deficiency (transplant etc.), neurologic disorder, and rheumatology disorder.
<sup>3</sup>There were no differences concerning ACE inhibitor, anticoagulants, insulin and other oral hypoglycemic agent users.

### Clinical Outcomes

Patients in cohort group A, treated with combination therapy have had better clinical outcomes. They had significantly lower hospitalization rate (5.8% combination, 22.3% control; *P = 0*.*0011*) and ER visit rate (7.8% combination, 23.3% control; *P = 0*.*0021*). The duration of disease was significantly less in combination group (6.67 ± 2.30 days combination, 18.71 ± 6.49 days control; *P < 0*.*0001*). Although, the control group had higher clinical score initially, the average scores in both cohort groups were greater than four, indicating higher risk of adverse outcomes comparably (35). The results from the Kaplan-Meier curve (**Figure 2**) and log-rank test show a significant difference in time to hospitalization, p-value < 0.001 between patients who received combination therapy compared to the patients who received standard control therapy. Thus, patients on the combination therapy had significantly lower probability of hospitalization compared to patients on the standard control therapy. In a multivariate analysis, where we adjusted for appropriate covariates, the odds ratio for hospitalization was significantly lower in the combination therapy group than the control therapy group (OR 0.177, CI 0.065-0.486; *P = 0*.*0008*), **Table 3**.

**Figure 2.**
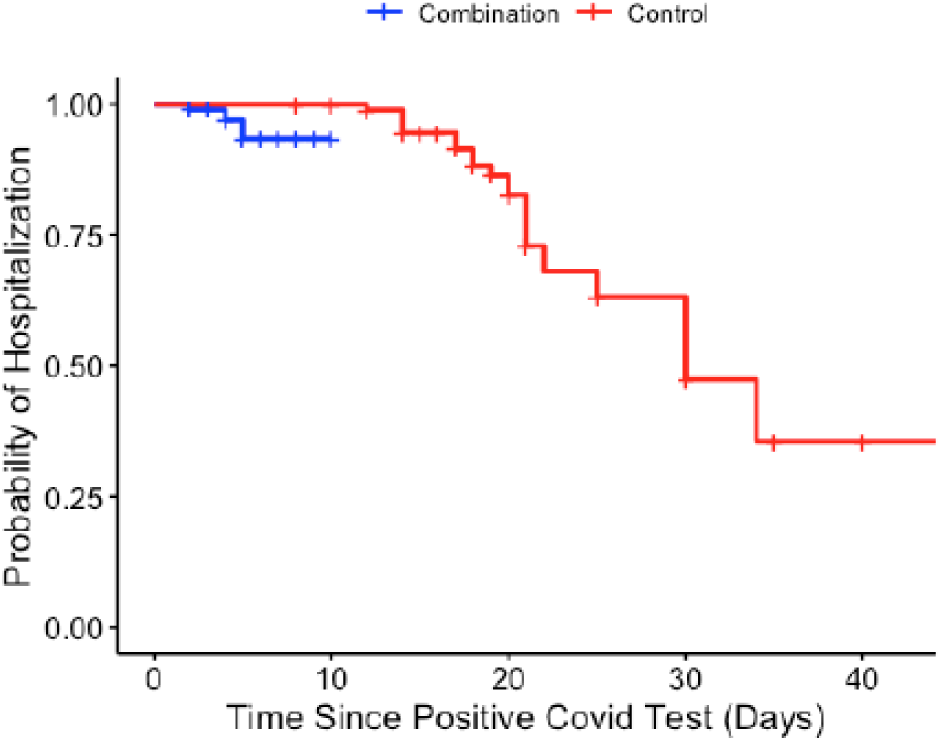
The Kaplan-Meier survival curves indicates the difference between the hospitalizations across the groups. The hospitalization curve for combination group was lower than control group.

**Table 3.** Odds ratio for hospitalization for outpatient COVID-19 cases

| *Variable* | *Odds ratio (95% confidence interval)* | *P-Value* |
| --- | --- | --- |
| *Age* | 1.026 (0.986-1.068) | 0.1987 |
| *Sex (female vs male)* | 0.437 (0.185-1.031) | 0.0588 |
| *Combination vs Control* | 0.177 (0.065-0.486) | 0.0008 |
| *Vaccination history, yes vs no* | 1.015 (0.302-3.410) | 0.9809 |
| *Previous history of COVID-19, yes vs no* | 0.637 (0.126-3.223) | 0.5859 |
| *Comorbidities<sup>1</sup>, yes vs no* | 3.104 (1.110-8.678) | 0.0308 |

<sup>1</sup>Comorbidities consist of diabetes, hypertension, cardiovascular disease, chronic kidney disease, chronic lung disease, chronic liver disease, hypothyroidism, rheumatologic disease, neurologic disease, immune deficiency (transplant etc.), hematologic disorder and cancer.

In terms of medication related complications, three patients in combination group reported adverse events between 2 and 6 days after starting medications, two reported drops in blood pressure and blood sugar and one indicated drops in blood pressure. However, they did not need an ER visit or hospitalization and recovered after stopping medications.

## Discussion

The COVID-19 pandemic continues to affect many countries even after vaccination drives and the presence of promising treatments, such as antivirals, convalescent plasma transfusion therapy and monoclonal antibodies (37). Vaccination was broadly advertised with the aim of reducing hospitalization and death, which was shown to be beneficial (3). However, there is a concern regarding their availability in developing countries (38) and efficacy against certain variants of SARS-CoV-2 strains (4). Furthermore, early treatment of outpatient cases with self-isolation at home seems to be helpful for reducing hospitalization and mortality effectively (39). Therefore, it is important to introduce an effective medication, which can be used safely for outpatients, to improve clinical outcomes and reduce hospitalization of COVID-19.

In the current study, the spironolactone and sitagliptin combination therapy reduced duration of illness, hospitalization rate, and ER visit rate significantly without causing serious adverse events. We now briefly review other existing therapies juxtaposed against the proposed combination therapy. The reduction of hospitalization rate with our combination therapy (74%) was better than that of inhaled corticosteroids (36%) (40) and molnupiravir (51%) (41). Although there are other medications such as nirmatrelvir (89%) (42), remdesivir (86%) (43), Paxlovid (88%) (44), and the monoclonal antibodies (70 – 85%) (7,45-47) with slightly better outcomes, the results may not be directly comparable. For example, on average, our patients are sicker than the patients treated with nirmatrelvir, remdesivir, and Paxlovid; this is based on maximum number of days elapsed since symptom onset for eligibility to be part of trials for remdesivir (within 7 days) (43), nirmatrelvir (within 3 days) (42), and Paxlovid (within 5 days) (44). The patients in our trial had an average onset duration of 6.8 days with several being seen two weeks after onset. Furthermore, all these antiviral drugs may typically be causing side effects, especially for patients with liver disease and CKD (48,49), whereas our combination therapy does not pose such major risks. It is also important to note that monoclonal antibodies and Paxlovid are not widely available in most parts of the world, especially in low- and middle-income countries (50). The authors of this paper are from Iran, Ghana, and India and as of now Paxlovid is not available in these countries. In case of monoclonal antibodies, recurrent mutations that lead to new variants of SARS-CoV-2 can jeopardize their efficacy (10,11,51). Overall, the ease of access, lower cost, fewer side effects and lack of significant drug interactions are the potential advantages of the combination therapy over the monoclonal antibodies and antivirals (52).

The proposed mechanisms of the tested combination therapy for improved outcomes include blocking virus entry, reducing viral replication, anti-inflammatory, antithrombotic, and antifibrotic effects by alleviating virus mediated inflammation or oxidative injury (21,53-55). A reasonable mechanism for increased virus infectivity would be the changes in biophysics of virus after attaching to soluble ACE2, which increases the weight and radius of viral particles, and results in escalating virus entry into the human cells (19,55). It has been shown that administration of the bioengineered soluble ACE2 variant, which was shorter than native soluble ACE2, could effectively reduce the infectivity of SARS-CoV-2 (56,57). This could be due to two reasons: (1) soluble ACE2 acting as a decoy receptor to reduce viral uptake or (2) native soluble ACE2 being replaced by the lighter recombinant soluble ACE2, resulting in lighter viral particles with lowered ability for cell entry. TMPRSS2 and ADAM17 (TNF-alpha converting enzyme) cleave ACE2 and potentiate viral uptake (58). The SARS-CoV-2 infection increases expression of ADAM17 and TMPRSS2 (18), down regulates ACE2 receptor. The activation of shedding process of ACE2 can potentially trigger inflammation (59,60). Therefore, SARS-CoV-2 infection initiates a vicious cycle of increasing soluble ACE2 level, more infectivity, reducing ACE2 on cell membrane and further inflammation. Spironolactone has negative effects on TMPRSS-2 (20,21) and ADAM17 (22), which leads to breaking the vicious cycle and reducing infectivity of SARS-CoV-2. Accordingly, the benefits of spironolactone on clinical outcomes of COVID-19 cases have been reported (61,62).

However, the replication of SARS-CoV-2 during the acute infection may overwhelm the ACE2 receptor, in which results in attachment of virus to other possible receptors, such as DPP4 (CD26), CD147, neurophilin 1, lectins, CD209L, and Tyrosine-protein kinase receptor UFO (AXL) (14,63-67). The use of DPP4 inhibitor was associated with lower mortality in diabetic COVID-19 patients (25,27,68), and even could improve the clinical end points and inflammation in non-diabetic COVID-19 cases (28). The proposed mechanisms could be the immunomodulatory functions (29), reduction of virus entry and diminishing viral replication. Therefore, the combination of DPP4 inhibitors and spironolactone could potentially improve the clinical outcomes better than each one alone (36).

Sitagliptin, a DPP4 inhibitor, and spironolactone are relatively safe medications without significant adverse side effects (69-71) or crossed side effects. Therefore, the combination of sitagliptin and spironolactone would be a relatively safe approach. It can potentially improve the clinical outcomes of COVID-19 cases without increasing risk of adverse side effects. In the current study, three patients (2.9%) reported medication related adverse side effects, such as hypotension and hypoglycemia, which were tolerable. The complications of spironolactone, (increased diuresis and orthostatic hypotension (71)) and sitagliptin (hypoglycemia (70)) can be managed by lowering the dose.

There are some limitations in the current study, including not having a double-blind design with considerably greater sample size and statistically significant difference in clinical score between control and treatment groups at initial presentation. The difference in clinical score on the first day might be less important; because of the given range of 0-29, the difference of 2 points in the severity score may not be as critical; both groups average above 4 and hence are deemed comparable in terms of adverse outcome potential (35). Another potential limitation is that our cohort includes 50% probable SARS-CoV-2 infection cases that have not had a PCR test-based confirmation. We had to follow the CDC guidelines (32) to treat probable cases with the proposed combination therapy as it would have been unethical to deliberately exclude them from the study. However, we analyzed the clinical score, time from symptoms onset, ER visit, hospitalization rate and duration of disease among PCR+ patients separately. The results were almost similar to the full patient group in our study (**Table 2**). Furthermore, almost half of the patients (57.3% control, and 49.5% combination) had COVID-19 positive PCR results, and others had history of exposure to COVID-19 with typical manifestations. We assumed that recall bias and loss to follow-up are distributed evenly across the two cohort groups.

**In conclusion**, combination of sitagliptin and spironolactone can reduce hospitalization, ER visits and duration of disease in acutely ill outpatient COVID-19 cases significantly. However, running a double blind, randomized, placebo-controlled trial is required to further substantiate the beneficial effects of this combination therapy in COVID-19 patients.

## Data Availability

The data produced in the present study are available upon reasonable request to authors.

## Abbreviations

ACE2: angiotensin-converting enzyme 2
ADAM17: Disintegrin and metalloproteinase domain-containing protein 17
COVID-19: coronavirus disease 2019
DPP4: dipeptidyl peptidase-4
ER: emergency room
SARS-CoV-2: severe acute respiratory syndrome coronavirus 2
SUMS: Shiraz University of Medical Sciences
TMPRSS2: transmembrane protease serine 2

## Acknowledgments

The authors express their gratitude to the Shiraz University Clinics for helping with data collection.

## Iranian Registry of Clinical Trial

IRCT registration number: IRCT20201003048904N2, Registration date: December 10, 2020

## Authors’ contributions

KA proposed the idea and designed the study. KA, RA, YM and RK wrote the manuscript. MAD, YM, HF, FSRR, AP, and MH collected the data. RA provided statistical analysis and wrote statistical part. All authors approved the final version of the manuscript.

## Data Availability

Shiraz University (MAD, YM, HF, FSRR, AP and MH) generated the data. University of Kentucky (KA and RA) has de-identified data and analyzed (RA) the data. The de-identified data are available for further investigations.

